# Safety and efficacy of the portable steam sauna bath for fluid removal in maintenance haemodialysis patients: A pilot phase II clinical trial

**DOI:** 10.1101/2020.06.21.20136648

**Authors:** Niranjini Chandrashekaran, Anna T. Valson, Ankita Priya, Sathya Subramani

**Affiliations:** Department of Physiology, Christian Medical College, Vellore, Tamil Nadu – 632004, India; Department of Nephrology, Christian Medical College, Vellore, Tamil Nadu – 632004, India

**Author notes:** **Corresponding Author: Name:** Anna T Valson, **Address:** Department of Nephrology, Christian Medical College, Vellore, Tamil Nadu – 632004, India, **Facsimile numbers**, **E-mail address:**.

**Keywords:** *Hammam*, haemodialysis, steam sauna, sweating, volume overload

## Abstract

**Background:** Dry heat, immersive, and *Hammam* sauna baths have been shown to aid fluid removal in haemodialysis patients but require high ambient temperatures, large volumes of water and sufficient space, all of which limit their widespread use in India. We aimed to study the safety and efficacy of a commercially available, inexpensive, portable steam sauna bath for this purpose.

**Methods:** In this pilot phase II clinical trial, six adult prevalent haemodialysis patients each underwent 6 sauna sessions lasting 30-60 minutes, on all non-dialysis days, for 2 weeks. Weight, blood pressure, serum urea, creatinine, electrolytes, haematocrit, core body temperature, thermal comfort, and thirst visual analogue scale were measured before and after each session. Karnofsky performance status (KPS) and Dialysis Symptoms Index (DSI) were measured at the beginning and end of the intervention period. The primary end points were per session weight loss and interdialytic weight gain (IDWG).

**Results:** Patients experienced a median weight loss of 0.35 kg, median fall in systolic and diastolic BP of 10 mm Hg and 2 mm Hg respectively (p < 0.001 for all) without significant change in IDWG (p = 0.46). Mean thermal comfort was 5.41 ± 0.56 out of 8, there was no significant increase in thirst (p = 0.06) and no significant change in KPS and DSI scores (p = 1.00 and 0.32 respectively). No adverse events were noted.

**Conclusions:** The portable steam sauna is safe, but modestly effective for fluid removal in haemodialysis patients, and may not influence IDWG.

**Details of each author’s contributions:** 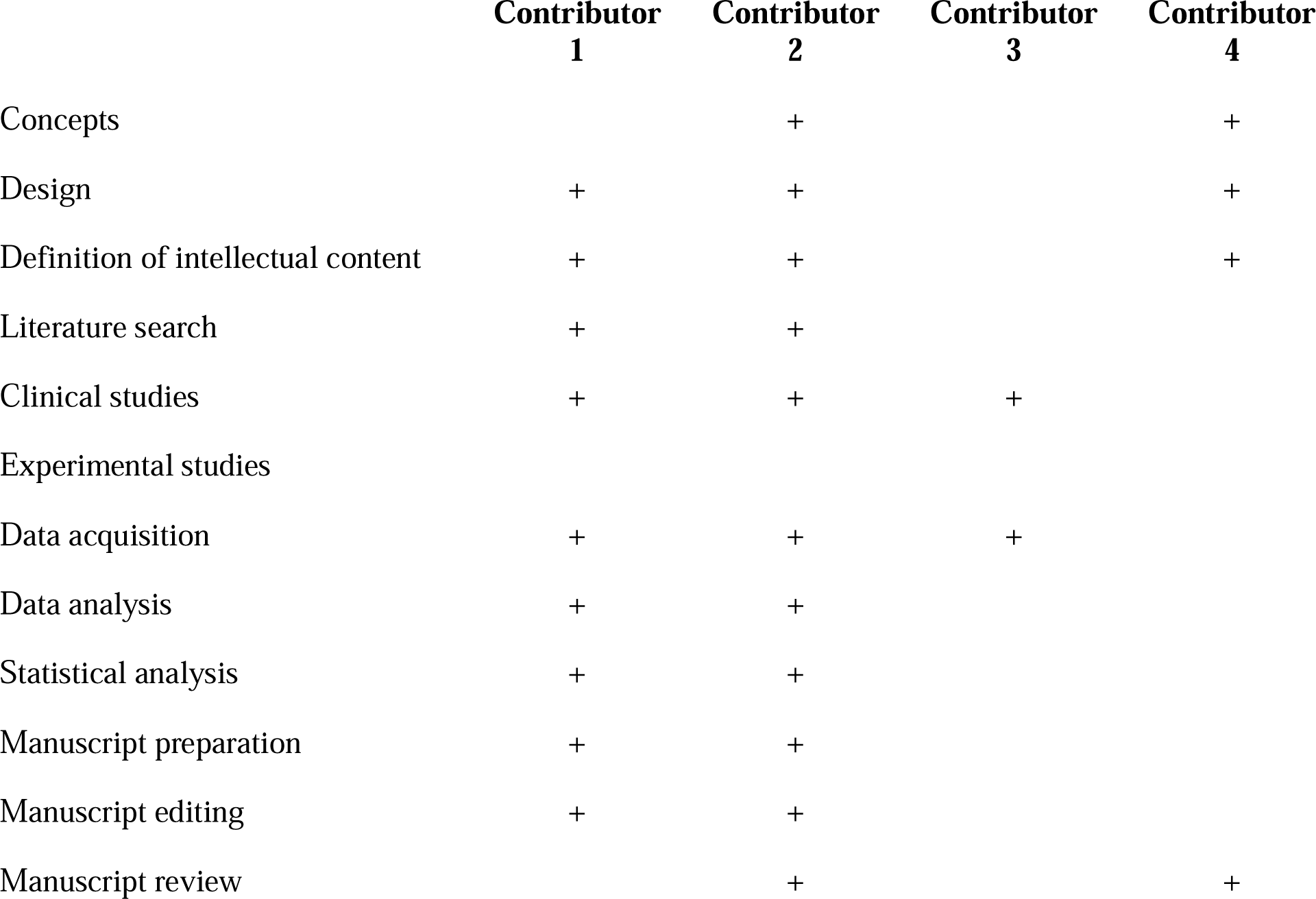

## INTRODUCTION

Volume overload in haemodialysis (HD) patients is managed by salt and fluid restriction[1], increasing the frequency[2,3] and/or duration[4] of HD sessions. These interventions add to the cost of treatment for self-paying patients, and the workload of busy dialysis units.

Sauna baths are a novel adjunct to HD for fluid removal. Three types of sauna therapy have been studied for fluid management in HD patients. The dry air sauna, also known as the Finnish bath, consists of a wood-panelled room measuring at least 3 m^2^ with an ambient air temperature of 80 - 100°C and humidity of 10-20%[5]. The average session consists of 2 ‘hot stays’ lasting 15 minutes each, interspersed with 10-minute cooling off periods, resulting in a mean weight loss of 780 ± 130 g per session[6]. The Turkish *hammam* or steam bath consists of a marble or ceramic tiled room with an ambient temperature of 37-43°C and 80% humidity, in which patrons spend 30 minutes, followed by a cooling off room where they splash themselves with cold water[7]. The weight loss achieved in this method is reported to be 420 ± 400 g per session. The hot water sauna bath involves immersion up to the neck for 30 minutes, in water maintained at a constant temperature of 37 - 43°C, with weight loss of 600 ± 700 g per session[7, 8].

Unlike Scandinavia, Turkey, and Japan, sauna use is not common in India, for various reasons. Because of the ambient temperatures in the long tropical summer season and the risk of heat stroke, there are safety concerns with the dry air sauna. Immersive hot water saunas require a minimum of 20 litres of water per session, which is impractical in water-scarce countries. While the Turkish *hammam* bath appears most suitable, space and water requirements are deterrents. In this context, we studied the safety and efficacy of a commercially available portable steam sauna bath for fluid removal in HD patients, which would offer the advantages of minimal water requirement, compactness, safe ambient temperature, and low input and running costs, besides portability.

## METHODS

This pilot Phase II interventional clinical trial was carried out to evaluate the safety and efficacy of a portable steam sauna bath as an adjunct therapy for fluid removal in maintenance HD patients. The sauna bath can be purchased via e-commerce portals in India, and costs approximately 70 USD. It consisted of a kettle in which 500 ml of water was boiled to produce steam which was conveyed via attached tubing into a tent within which the patient was seated, head-out (Figure 1).

**Figure 1:**
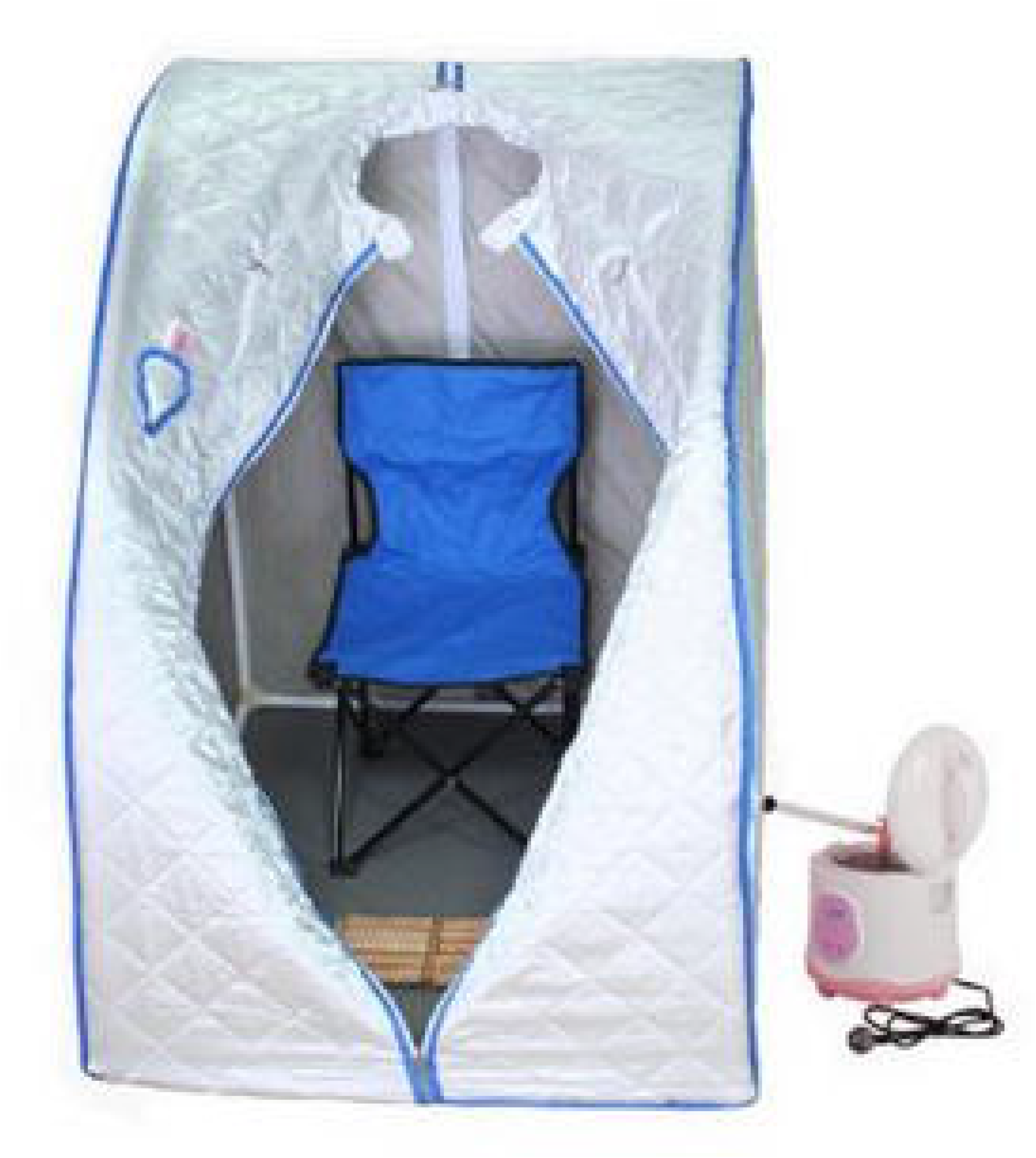
Sauna apparatus used for the study.

The protocol was approved by the Institutional Review Board and Ethics Committee of our institution. Written, informed consent was obtained from all participants. The trial was registered with the Clinical Trials Registry of India (CTRI/2017/02/007823) and overseen by the institution’s data safety monitoring board (DSMB).

### Objectives

The primary objective was to study the effect of the portable steam sauna on the volume status of prevalent HD patients, as measured by difference in weight before and after a sauna session and mean interdialytic weight gain (IDWG) during the intervention period.

The secondary objectives were to study the effect of sauna baths on blood pressure, serum electrolytes, urea, creatinine, haematocrit, core body temperature, thirst, thermal comfort and quality of life, besides documenting any adverse events that might occur in patients undergoing this therapy.

### Inclusion criteria

Adult (≥ 18 years) prevalent CKD Stage G5D patients were invited to participate in the study if they were on thrice weekly or twice weekly HD for ≥ three months, had a well-functioning arteriovenous fistula with no vascular access intervention in the preceding three months, had a mean IDWG ≥ 1.5 kg over a two-week observation period and no major illness requiring hospitalization in the preceding one month. We excluded patients who were unable or unwilling to provide informed consent, patients with any cardiovascular event during the 12 months preceding this study (myocardial infarct, heart failure [New York Heart Association stage III/IV], heart rhythm disorders, cerebrovascular accident, stable or unstable angina, symptomatic and/or operated peripheral vascular disease), patients with past history of heat stroke, patients in sepsis or with open surgical wounds (including HD catheters).

The pre-intervention observation phase, recruitment, trial session, intervention phase and post-intervention observation phase were completed between April 1^st^ –July 12^th^ 2017.

### Pre-intervention observation phase and trial session

The study flow diagram as per STROBE guidelines is given in Figure 2. All patients in the dialysis unit (n = 180) were observed for two weeks to assess their baseline clinical status, average IDWG, blood pressure and vascular access. Patients who fulfilled the inclusion criteria were invited to participate in the study (n = 65). In those who consented to participate (n = 6), a single trial session was offered during which the patients were made to sit in the sauna bath unit to establish thermal comfort. If they tolerated the procedure well with no significant discomfort or adverse events, they were recruited to take part in the study.

**Figure 2:**
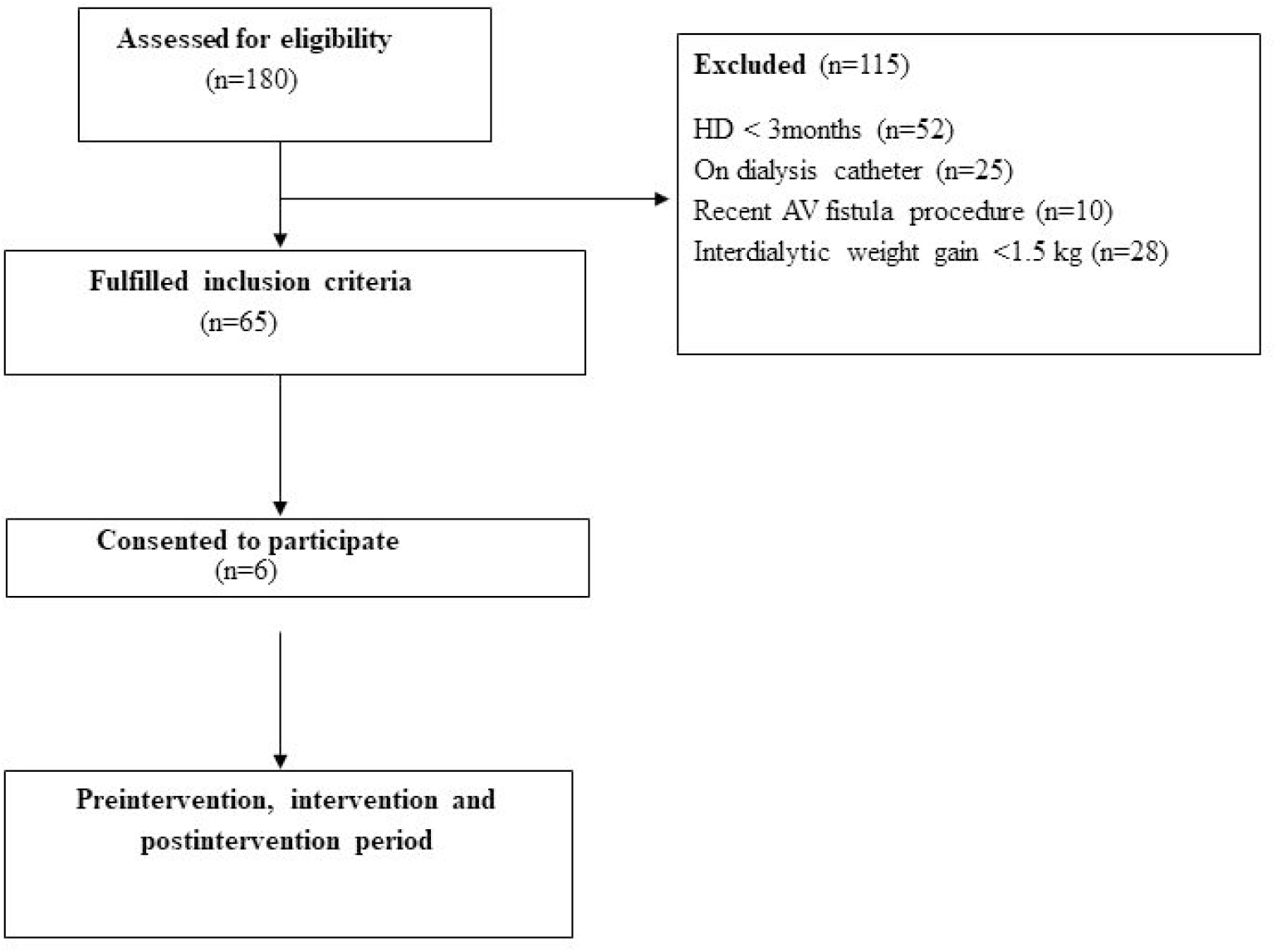
Study flow diagram.

### Intervention phase

The intervention period lasted two weeks and the study setting was the dialysis unit of our institution. During this period, timing and duration of HD sessions, medication doses, and dry weight remained unchanged. Patients were asked to come to the dialysis unit on their non-dialysis days, thrice weekly and sit (head out) in a commercially available portable steam sauna bath unit measuring 48cm ⨯ 33cm ⨯ 48cm (Kawachi, Mumbai, India) at 40□ C and 100% humidity for 30-60 mins (as tolerated) for a total of 6 sessions per participant. Prior to each sauna bath, the patients and volunteers were advised to have a warm shower and scrub their skin thoroughly to aid in unclogging their sweat pores. Meals or heavy exercise were not allowed in the one hour before and two hours after each session. On the days of the intervention, the patients were advised not to take their morning dose of antihypertensive medication to prevent excessive fall of blood pressure during the session and avoid confounding. Being a pragmatic clinical trial, we did not ask patients to restrict their fluid intake during the intervention phase.

### Measurements

Body weight was measured before and after each session (after complete drying) using a standardised and calibrated electronic weighing machine. Body surface area was calculated using DuBois formula[9], given by BSA = 0.007184 * Height^0.725^ * Weight^0.425^ where height is in centimeters and weight in kilograms. Sweat rate was expressed as kg/BSA/min since sweat gland density is inversely proportional to BSA[10].

Core body temperature was taken as the oral temperature measured by a digital thermometer before and after each session. Blood pressure was measured before, during (every 15 minutes) and after each session using an aneroid sphygmomanometer. Serum sodium, potassium, chloride, ionized calcium, urea, creatinine and haematocrit were assessed before and after each session on venous samples taken under aseptic precautions and analysed by a point-of-care portable blood analyser (*i-STAT* handheld, Abbot Point of Care Inc. Princeton, NJ, USA) which is able to measure multiple parameters in the same patient using 2-3 drops of blood fed into a single-use multi-sensor cartridge provided by the manufacturer. Creatine phosphokinase (CPK) levels were measured before and after the second sauna session of the week, once weekly, with the assay being carried out by the clinical biochemistry laboratory.

Modified Bedford thermal comfort scale (MBTCS) is a 7 point scale that measures the thermal comfort experienced by a person and was assessed midway through each session[11]. Thirst intensity during each session was assessed using a 10 cm thirst visual analogue scale with 0 for *no thirst* and 10 for *worst possible thirst*[12]. Quality of life was assessed using the Karnofsky Performance Status Scale (KPS)[13] and Dialysis Symptoms Index(DSI)[14] at the start and end of the intervention period.

### Post-intervention observation phase

After the end of the intervention phase, each patient entered a two-week observation phase when sauna sessions were discontinued and patients continued with their regular HD schedule. Their IDWG and pre-dialysis systolic and diastolic BP were monitored during this period to determine any residual effect of the intervention phase.

### Comparison with healthy volunteers

Nine healthy volunteers undertook a single sauna session lasting 30 minutes in the same apparatus, prior to this trial. The effect of sauna baths on HD patients and these healthy volunteers was compared.

### Statistical analysis

Statistical analysis was carried out using Statistical Package for Social Sciences v. 16.0 (SPSS Inc. Chicago, USA). This being a pilot study, no power calculation was done. Quantitative variables were expressed using mean ± standard deviation (for normally distributed variables) or median with inter-quartile range (for variables with a skewed distribution). Students’ paired T Test or Wilcoxon signed rank test was used as appropriate to compare pre- and post intervention parameters. A p value < 0.05 was considered significant.

## RESULTS

Six HD patients (five of whom were male) volunteered to participate in the trial. Table 1 shows their baseline demographic and clinical details.

**Table 1:**
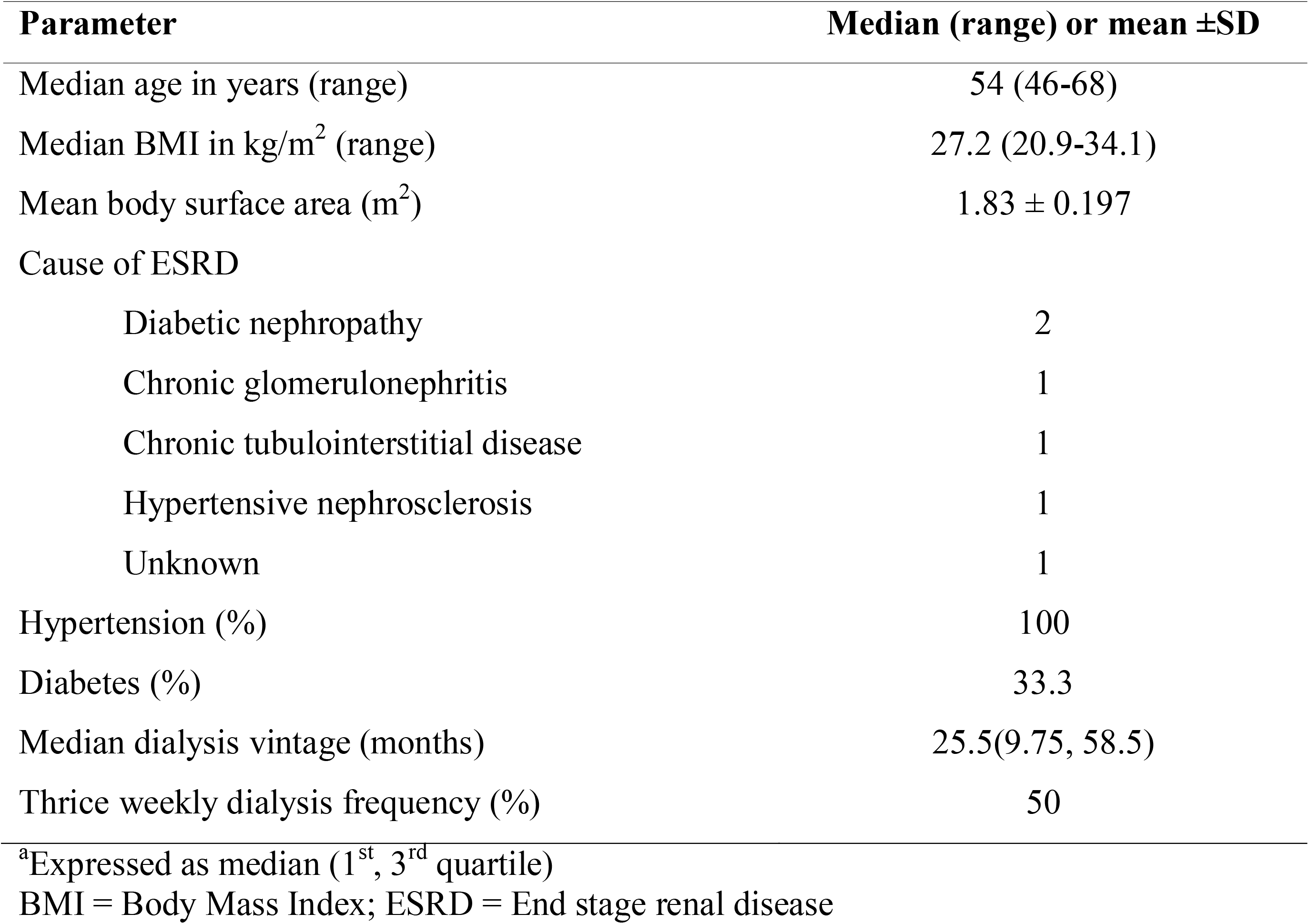
Demographic and clinical details of the study patients (n = 6)

| **Parameter** | **Median (range) or mean ±SD** |
| --- | --- |
| Median age in years (range) | 54 (46-68) |
| Median BMI in kg/m<sup>2</sup> (range) | 27.2 (20.9-34.1) |
| Mean body surface area (m<sup>2</sup>) | 1.83 ± 0.197 |
| Cause of ESRD |  |
| Diabetic nephropathy | 2 |
| Chronic glomerulonephritis | 1 |
| Chronic tubulointerstitial disease | 1 |
| Hypertensive nephrosclerosis | 1 |
| Unknown | 1 |
| Hypertension (%) | 100 |
| Diabetes (%) | 33.3 |
| Median dialysis vintage (months) | 25.5(9.75, 58.5) |
| Thrice weekly dialysis frequency (%) | 50 |

<sup>a</sup>Expressed as median (1<sup>st</sup>, 3<sup>rd</sup> quartile)
BMI = Body Mass Index; ESRD = End stage renal disease

None of the patients were on ACE inhibitors, angiotensin receptor blockers or mineralocorticoid receptor antagonists, which can potentially affect potassium excretion by the kidney and sweat glands.

Table 2 shows the effect of 6 steam sauna sessions on various physiological, laboratory, and quality of life parameters for each individual study patient and the group as a whole. Mean duration of each session was 45.39 ± 11.17 minutes and mean per session weight loss per unit body surface area (BSA) was 0.0047 kg/m^2^ BSA/min. The following parameters showed a significant change after sauna sessions: pulse rate (+3.2/min, p = 0.009), respiratory rate (+1.8/min, p < 0.001), core temperature (+ 1°C, p< 0.001), median weight (−0.35 kg, p < 0.001), serum creatinine (+0.5 mg/dL, p < 0.001), systolic blood pressure (−10 mm Hg, p < 0.001), diastolic blood pressure (−2 mm Hg, p = 0.001), serum chloride (+1.9 meq/L, p=0.006). Mean MBTCS midway through each session was 5.41 ± 0.56, where 8 was the maximum score. There was no significant change in KPS or DSI scores, and no adverse events were observed during the intervention period. DSI scores for individual patients across all components are given in Table 3.

**Table 2:**
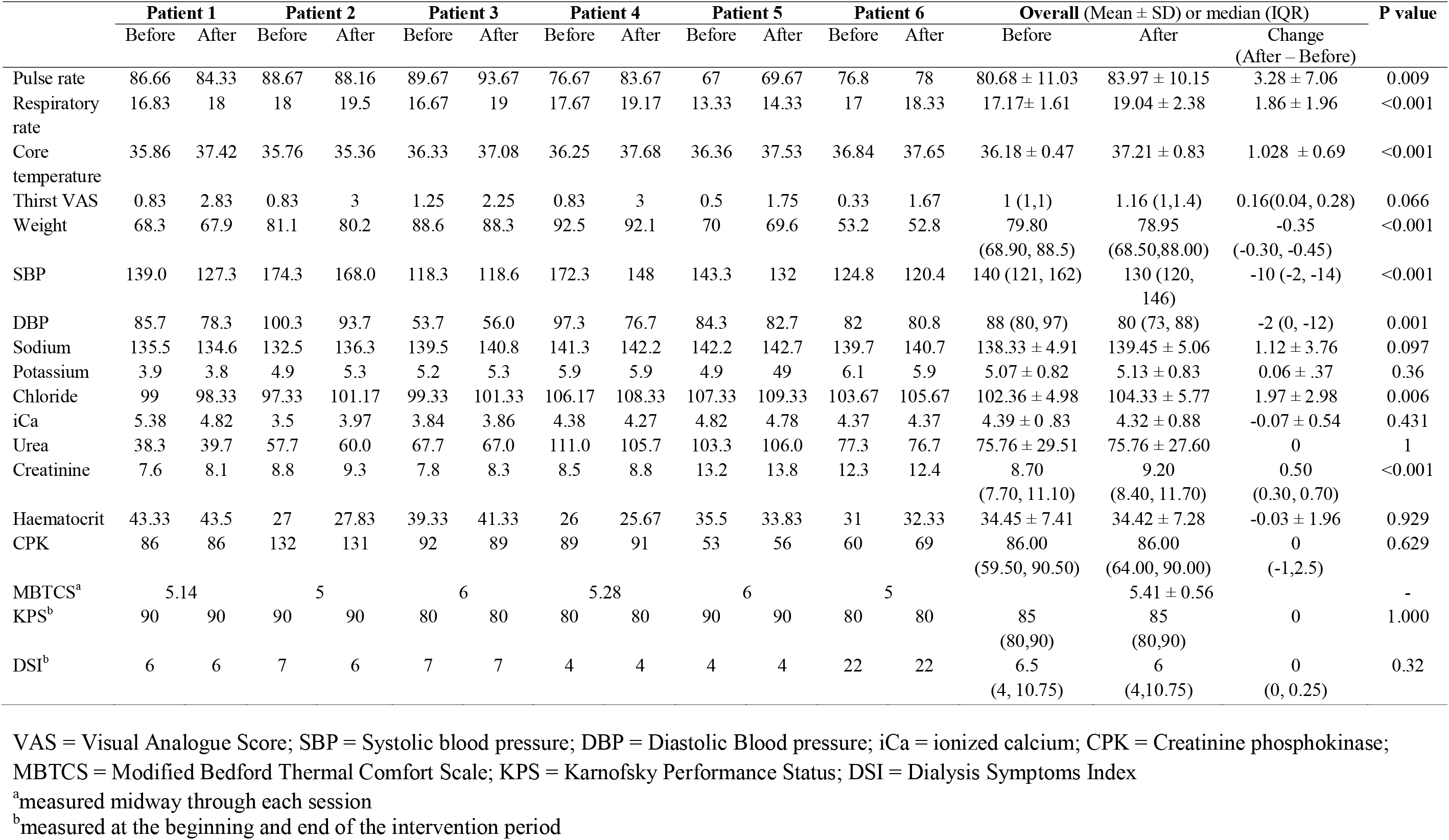
Effect of steam sauna on various physiological parameters in HD patients (n = 6, average of 6 sessions).

**Table 3:** Dialysis Symptom Index score for individual patients before and after the intervention period across various components.

| S. No | Symptom | Patient 1 |  | Patient 2 |  | Patient 3 |  | Patient 4 |  | Patient 5 |  | Patient 6 |  |
| --- | --- | --- | --- | --- | --- | --- | --- | --- | --- | --- | --- | --- | --- |
|  |  | Before | After | Before | After | Before | After | Before | After | Before | After | Before | After |
| 1 | Constipation | 0 | 0 | 0 | 0 | 0 | 0 | 0 | 0 | 0 | 0 | 0 | 0 |
| 2 | Nausea | 0 | 0 | 2 | 2 | 0 | 0 | 0 | 0 | 0 | 0 | 2 | 2 |
| 3 | Vomiting | 0 | 0 | 0 | 0 | 0 | 0 | 0 | 0 | 0 | 0 | 0 | 0 |
| 4 | Diarrhea | 0 | 0 | 0 | 0 | 0 | 0 | 0 | 0 | 0 | 0 | 0 | 0 |
| 5 | Decreased appetite | 0 | 0 | 2 | 2 | 0 | 0 | 2 | 2 | 0 | 0 | 2 | 2 |
| 6 | Muscle Cramps | 0 | 0 | 0 | 0 | 0 | 0 | 0 | 0 | 0 | 0 | 0 | 0 |
| 7 | Swelling in legs | 0 | 0 | 0 | 0 | 1 | 1 | 0 | 0 | 1 | 1 | 1 | 1 |
| 8 | Shortness of breath | 2 | 2 | 3 | 2 | 2 | 2 | 0 | 0 | 2 | 2 | 3 | 3 |
| 9 | Dizziness | 0 | 0 | 0 | 0 | 0 | 0 | 0 | 0 | 0 | 0 | 0 | 0 |
| 10 | Restless legs | 0 | 0 | 0 | 0 | 0 | 0 | 0 | 0 | 0 | 0 | 0 | 0 |
| 11 | Numbness/ Tingling in feet | 0 | 0 | 0 | 0 | 0 | 0 | 0 | 0 | 0 | 0 | 0 | 0 |
| 12 | Feeling tired | 2 | 2 | 0 | 0 | 2 | 2 | 2 | 2 | 1 | 1 | 4 | 4 |
| 13 | Cough | 0 | 0 | 0 | 0 | 0 | 0 | 0 | 0 | 0 | 0 | 2 | 2 |
| 14 | Dry mouth | 0 | 0 | 0 | 0 | 0 | 0 | 0 | 0 | 0 | 0 | 0 | 0 |
| 15 | Bone/Joint pain | 0 | 0 | 0 | 0 | 1 | 1 | 0 | 0 | 0 | 0 | 0 | 0 |
| 16 | Chest pain | 0 | 0 | 0 | 0 | 0 | 0 | 0 | 0 | 0 | 0 | 0 | 0 |
| 17 | Headache | 0 | 0 | 0 | 0 | 0 | 0 | 0 | 0 | 0 | 0 | 2 | 2 |
| 18 | Muscle soreness | 0 | 0 | 0 | 0 | 0 | 0 | 0 | 0 | 0 | 0 | 0 | 0 |
| 19 | Difficulty concentrating | 0 | 0 | 0 | 0 | 0 | 0 | 0 | 0 | 0 | 0 | 0 | 0 |
| 20 | Dry skin | 0 | 0 | 0 | 0 | 0 | 0 | 0 | 0 | 0 | 0 | 2 | 2 |
| 21 | Itching | 0 | 0 | 0 | 0 | 0 | 0 | 0 | 0 | 0 | 0 | 0 | 0 |
| 22 | Worrying | 0 | 0 | 0 | 0 | 1 | 1 | 0 | 0 | 0 | 0 | 2 | 2 |
| 23 | Feeling nervous | 0 | 0 | 0 | 0 | 0 | 0 | 0 | 0 | 0 | 0 | 0 | 0 |
| 24 | Trouble falling asleep | 2 | 2 | 0 | 0 | 0 | 0 | 0 | 0 | 0 | 0 | 2 | 2 |
| 25 | Trouble staying asleep | 0 | 0 | 0 | 0 | 0 | 0 | 0 | 0 | 0 | 0 | 0 | 0 |
| 26 | Feeling irritable | 0 | 0 | 0 | 0 | 0 | 0 | 0 | 0 | 0 | 0 | 0 | 0 |
| 27 | Feeling sad | 0 | 0 | 0 | 0 | 0 | 0 | 0 | 0 | 0 | 0 | 0 | 0 |
| 28 | Feeling anxious | 0 | 0 | 0 | 0 | 0 | 0 | 0 | 0 | 0 | 0 | 0 | 0 |
| 29 | Decreased interest in sex | 0 | 0 | 0 | 0 | 0 | 0 | 0 | 0 | 0 | 0 | 0 | 0 |
| 30 | Difficulty being sexually aroused | 0 | 0 | 0 | 0 | 0 | 0 | 0 | 0 | 0 | 0 | 0 | 0 |
| Total |  | 6 | 6 | 7 | 6 | 7 | 7 | 4 | 4 | 4 | 4 | 22 | 22 |

Table 4 details the changes in IDWG and pre-dialysis systolic and diastolic BP through the pre-intervention observation period, intervention period and post-intervention observation period for individual patients and the group as a whole. Although the median IDWG decreased from 3.0 kg to 2.7 kg in from the pre-intervention observation period to the intervention period, this was not statistically significant.

**Table 4:** Interdialytic weight gain and blood pressure in individual HD patients in the pre-intervention observation period (P1), intervention period (P2) and post-intervention observation period (P3)

|  | **IDWG**<br>**(kg)** | **Pre-dialysis SBP**<br>**(mm Hg)** | **Pre-dialysis DBP**<br>**(mm Hg)** |
| --- | --- | --- | --- |
| **Patient 1** |  |  |  |
| **P1** | 3.0 | 136.6 | 81.6 |
| **P2** | 2.6 | 140.0 | 82.0 |
| **P3** | 3.7 | 155.7 | 80.0 |
| **Patient 2** |  |  |  |
| **P1** | 3.3 | 168.3 | 90.0 |
| **P2** | 2.6 | 168.6 | 88.6 |
| **P3** | 3.8 | 181.4 | 95.7 |
| **Patient 3** |  |  |  |
| **P1** | 2.8 | 145.0 | 80.0 |
| **P2** | 2.4 | 133.3 | 80.0 |
| **P3** | 2.4 | 130.0 | 75.7 |
| **Patient 4** |  |  |  |
| **P1** | 3.0 | 161.6 | 93.3 |
| **P2** | 3.4 | 160.0 | 94.0 |
| **P3** | 3.1 | 147.1 | 82.8 |
| **Patient 5** |  |  |  |
| **P1** | 3.7 | 146.0 | 82.0 |
| **P2** | 4.2 | 140.0 | 82.0 |
| **P3** | 3.4 | 140.0 | 81.4 |
| **Patient 6** |  |  |  |
| **P1** | 2.4 | 128.0 | 80.0 |
| **P2** | 2.7 | 134.0 | 82.0 |
| **P3** | 2.3 | 116.0 | 77.1 |
| **Overall** |  |  |  |
| **P1, median (IQR)<sup>a</sup>** | 3.0<br>(2.8, 3.3) | 145.5<br>(138.7,157.7) | 81.80<br>(80.00, 90.00) |
| **P2, median (IQR)<sup>a</sup>** | 2.7<br>(2.6, 3.3) | 140<br>(135.5,155.0) | 82.00<br>(82.00, 86.93) |
| **P3, median (IQR)<sup>a</sup>** | 3.26<br>(2.38, 3.70) | 143.57<br>(130.00,155.70) | 80.72<br>(77.14, 82.8) |

IDWG: Inter-dialytic weight gain; SBP: Systolic blood pressure; DBP: Diastolic blood pressure
All values are average of 6 sessions unless specified
<sup>a</sup>p = not significant for P1 vs. P2, P2 vs. P3 and P1 vs. P3 for all parameters

There was a weakly positive correlation between per session weight loss and IDWG in the pre-intervention observation period (r = 0.299, p=0.565), weakly negative correlation between per session weight loss and IDWG in the intervention period (r= −0.11, p=0.536) and a weakly positive correlation between per session weight loss and IDWG in the post-intervention observation period (r= 0.264, p=0.119). None of these correlations were significant.

None of the patients experienced intradialytic hypotension during the pre-intervention observation period, intervention period or post-intervention observation period. In the post-intervention observation period, despite IDWG returning to pre-intervention levels, both median systolic and diastolic blood pressures showed a trend towards lower values, though again, this change was not statistically significant.

Table 5 compares HD patients and healthy volunteers with respect to demographic parameters and physiological effects of sauna. Healthy volunteers were younger, and had significantly greater weight loss per unit body surface area compared to HD patients.

**Table 5:** Comparison between healthy volunteers and study patients with regard to demographic characteristics and per session (post– pre session) change in parameters.

|  | **Healthy**<br>**volunteers (n=9)** | **HD patients**<br>**(n=6)** | **P value** |
| --- | --- | --- | --- |
| **Age in years** | 29(26-38) | 54 (46-68) | < 0.001 |
| **Gender (male)** | 44.4% | 83.3% | 0.132 |
| **Mean BSA (m<sup>2</sup>)** | 1.69 ± 0.17 | 1.83 ± 0.19 | 0.159 |
| **Median SBP change**<br>**per session (mm Hg)** | -4<br>(-10, 2) | -10<br>(-2, -14) | 0.140 |
| **Median DBP change**<br>**per session (mm Hg)** | 0<br>(-14, 0) | -2<br>(-12, 0) | 0.567 |
| **Median Per session**<br>**change in serum**<br>**sodium (meq/L)** | 0<br>(-1, 1) | 1<br>(0,2) | 0.086 |
| **Median Per session**<br>**change in serum**<br>**potassium (meq/L)** | -0.2<br>(-0.4, 0.1) | 0.1<br>(-0.2,0.2) | 0.061 |
| **Median Per session**<br>**change in haematocrit**<br>**(g/dL)** | 1<br>(0, 2) | 0<br>(-1,1) | 0.236 |
| **Median weight**<br>**change per session**<br>**(kg)** | -0.5<br>(-0.30, -0.7) | -0.35<br>(-0.30, -0.45) | 0.348 |
| **Median Weight loss**<br>**in kg/m<sup>2</sup> BSA/minute** | 0.01<br>(0.006,0.011) | 0.0047<br>(0.0029, 0.0067) | <0.001 |

BSA = Body surface area; SBP = systolic blood pressure; DBP = Diastolic BP

## DISCUSSION

This trial aimed to study the efficacy and safety of a commercially available portable steam sauna bath for fluid removal in HD patients. In addition, the effect of sauna baths on various physiological, laboratory, and quality of life indices was also examined. In terms of primary efficacy endpoints, we found that there was a significant median weight loss per sauna session, but no significant difference in IDWG. The physiological effects of sauna included a significant increase in pulse rate, respiratory rate, core temperature, serum creatinine, and serum chloride, with a significant fall in systolic and diastolic blood pressure post session. There was no significant change in quality of life indices (KPI and DSI) over the two-week intervention period, and no significant difference in IDWG, pre-dialysis systolic or diastolic blood pressure between the pre-intervention, intervention and post-intervention periods. The procedure was safe, with no adverse events noted, and no instance of intradialytic hypotension during the intervention period. HD patients had a markedly lower weight loss per unit BSA compared to healthy volunteers.

The median weight loss of 350g for HD patients in our study, was statistically significant, but lower than that reported in a similar study that compared immersive hot water baths in Switzerland and *Hammam* baths in Tunisia for reducing IDWG in HD patients[6]. The per session weight loss for *Hammam* users in that study was 420g, despite the mean duration of baths being shorter than in our study (30 minutes vs. 45 minutes). There are several explanations for this. The intervention period for the latter study was longer (4 weeks vs. 2 weeks), and its subjects were habitual *Hammam* users. It is well known that acclimatization increases the efficiency of sweating, including sweat rate and potassium loss[15]. Nevertheless, the authors reported that immersive hot water sauna baths resulted in a higher mean weight loss (600g) than *Hammam* baths. The reason why steam baths appear to be less efficient at inducing sweating than dry heat saunas[16] and immersive sauna baths[7] has been attributed to their high ambient humidity of 80-100%, which reduces the vaporization ratio (ratio of evaporated to excreted sweat), leading to the accumulation of a layer of sweat over the skin surface, which impedes further sweat secretion from the sweat glands[16]. Even so, the average weight loss in our study varied from 0.3 kg – 1.7 kg, indicating that some patients may respond better to the steam sauna than others.

Although median IDWG decreased from 3 kg to 2.7 kg during the intervention period, this was not statistically significant. There are at least two explanations for this: firstly, the sample size was small, and the net effect on IDWG was affected by marked between-patient variation, with 3 patients recording a decrease in average IDWG ranging from 0.4-0.7 kg, while 3 reported an increase in IDWG ranging from 0.3-0.5 kg. Second, we hypothesize that increased fluid intake in the interdialytic period, which may at least partly explain the between-patient differences in IDWG, may have attenuated the net effect of sauna baths on IDWG, as has been reported by other investigators[7]. The latter can be attributed both to the high ambient temperatures prevalent in India during the hot summer months during which the study was conducted, and to the differential physiological response of the body to wet heat compared to dry heat. There is evidence that steam saunas result in higher heat stress and heat discomfort compared to dry heat saunas because the lower vaporization ratio results in less evaporative cooling, accumulation of heat, and rise in core temperature[16]. Although there was no significant increase in thirst VAS scores after sauna sessions in our trial, there was significant inter-individual variation in the perception of thirst, which may have influenced their fluid intake between sessions. The weakly positive, though non-significant correlation between per session weight loss and IDWG in the pre- and post-intervention observation periods indicates that patients who sweat more under normal circumstances may compensate through increased fluid intake, which has a bearing on their IDWG. Longer or more frequent steam sauna baths may be required to increase weight loss and sufficiently negate the effect of increased fluid intake.

The haemodynamic changes associated with sauna baths depend on their ambient temperature and duration. Higher temperatures cause an initial rise in systolic blood pressure as thermoregulatory mechanisms cause sympathetic activation, increase in heart rate, and consequently, cardiac output[16]. As the duration of bathing increases, a fall in systolic blood pressure becomes apparent, presumably due to volume depletion[17]. Diastolic blood pressure falls in steam sauna baths[16, 17] due to peripheral vasodilation, which is also part of the thermoregulatory response to heat. HD patients in this study showed a significant increase in respiratory rate, heart rate, core body temperature, and fall in systolic and diastolic blood during steam sauna sessions, which are all are part of the physiological response to sauna[5].

Serum concentrations of creatinine and chloride showed a significant increase during sessions while sodium showed an increasing trend which was not significant. Sodium, chloride and creatinine concentrations in sweat are lower than that in serum[13], and therefore the loss of hypotonic fluid in sweat can theoretically result in a rise in their serum concentrations.

Despite IDWG and systolic blood pressure returning to pre-intervention levels at the end of the study period, the diastolic blood pressure showed a small, though insignificant fall. This suggests that even over a short study period, sauna therapy presents the possibility of vascular remodelling and may improve endothelial function by lowering diastolic pressure[19].

When compared to healthy volunteers, HD patients had a markedly lower sweat rate per unit body surface area (0.0047 vs. 0.01 kg/m^2^ BSA/min, p < 0.001), and the median per session weight loss was 0.5 kg in healthy volunteers compared to 0.35 kg in HD patients. CKD patients have been shown to have sweat gland atrophy[20] and reduced sweating in response to pilocarpine iontophoresis, which is even more marked in patients on dialysis[21]. In addition, healthy volunteers were significantly younger, and heat-activated sweat gland density and flow decreases with age[22].

Our study has certain limitations. The sample size was small and the study population was heterogenous in terms of its response to steam sauna. Patients were required to come to the dialysis unit on non-dialysis days and we were unable to offer sauna therapy at home, because safety studies should be conducted in healthcare facilities to allow prompt intervention in case of adverse events. This severely limited the number of patients who were willing to participate in the study. Nevertheless, this pilot, hypothesis generating study can form the basis for larger and longer trials to study the utility of this or other sauna modalities in HD patients. A second limitation is that the duration of the intervention was short and it is likely that with a longer study period, heat acclimatization may have increased the efficiency of sweat gland function resulting in a higher sweat rate, lower sodium and higher potassium loss[15]. The study period was also too short to demonstrate any change in functional capacity or quality of life in HD patients taking part in the study. Thirdly, we did not restrict or monitor water intake in study participants, which may have blunted the effect of sauna on IDWG. However, this was done to ensure the study simulated real life conditions, and all patients reported their joy at being able to increase fluid intake in parallel with sauna sessions. We did not measure residual urine output in HD patients and hence cannot comment on the effect of steam sauna on residual kidney function. Many parameters, such as pulse rate, respiratory rate, core temperature, urea, creatinine, chloride, CPK, ionized calcium, thermal comfort, thirst, and quality of life, were not measured for the healthy volunteers, and we are unable to comment on whether these were different between the two groups. A longer pre-intervention trial period could have helped us identify patients with a higher sweat rate, who may have benefited more from sauna therapy. Lastly, in their present form, there may be inherent limitations to the utility of portable steam saunas with respect to the efficiency of induced sweating, consequent heat stress, and impact on IDWG. Design modifications may therefore be required before the portable sauna can be routinely offered to HD patients to increase fluid removal over and above HD. Alternatively, other sauna modalities, such as far infra-red sauna, which can induce sweating with lower heat stress[23], may need to be explored.

## CONCLUSION

Portable steam sauna baths are a safe and modestly effective adjunct therapy for fluid removal in HD patients, though the present study could not demonstrate a significant effect on IDWG. Studies exploring other sauna modalities or a modified version of this device in a larger patient population over a longer intervention period are required to explore the potential of this low-cost adjunct to HD.

## Data Availability

All patient data is available from the authors on request.

## Acknowledgements

The study was funded by an Internal Fluid Research Grant from our institution

